# 2019 Novel Coronavirus can be detected in urine, blood, anal swabs and oropharyngeal swabs samples

**DOI:** 10.1101/2020.02.21.20026179

**Authors:** Liang Peng, Jing Liu, Wenxiong Xu, Qiumin Luo, Keji Deng, Bingliang Lin, Zhiliang Gao

**Author notes:** Correspondence: Professor Zhiliang Gao,; or Professor Bingliang Lin,. Department of Infectious Diseases, Guangdong Key Laboratory of Liver Disease Research, The 3rd Affiliated Hospital of Sun Yat-sen University, Guangzhou 510630, Guangdong, China. These first authors contributed equally to this article (Liang Peng, Jing Liu, Wenxiong Xu, Qiumin Luo).

## Abstract

We tested samples collected from nine patients diagnosed with coronavirus disease 2019 (COVID-19). The virus was found in urine, blood, anal swabs and oropharyngeal swabs. It is the first time for SARS-CoV-2 found in urine, though no urinary irritation was found.

## Introduction

2019 Novel Coronavirus (SARS-CoV-2) infection outbroke and spread to the world from 2019. By January 28, 2020, a total of 4593 global confirmed cases, of which 4537 were from China, had been reported(*1*). On January 30, 2020, SARS-CoV-2 infection was declared as a global health emergency by World Health Organization (WHO). The mechanism of SARS-CoV-2 infection and organ invasion are unclear, which directly leads to difficulties and blindness in clinical diagnosis and treatment. We aim to detect SARS-CoV-2 nucleic acid from urine, blood, anal swab and oropharyngeal swab samples. We hope to provide evidences of the multiple organ invasion of the virus and figure out its relation with clinical manifestations.

## Methods

Nine patients confirmed diagnosed with SARS-CoV-2 infection(*2*) were included in this prospective study. Urine, blood, anal swabs and oropharyngeal swabs from enrolled patients were obtained and detected SARS-CoV-2 RNA level by quantitative real-time polymerase chain reaction (qRT-PCR). Patients’ demographic data and clinical characteristics were recorded. This study was approved by the institutional review board (IRB) of the Third Affiliated Hospital of Sun Yat-sen University. All patients voluntarily signed an informed consent form approved by the IRB before participation.

## Results

Patient 7, a 31 years old female without any urinary irritation, had positive results of SARS-CoV-2 in both urine and oropharyngeal swab on the 7^th^ day after symptom onset. Patient 1 and 5 had negative results in oropharyngeal swab, on the 10^th^ and 15^th^ day after onset. Patient 8 had three positive results in blood, anal swab and oropharyngeal swab on the 3^rd^ day after onset. Viral load was roughly higher in anal swab than oropharyngeal swab in patient 9 (Table).

**Table.**
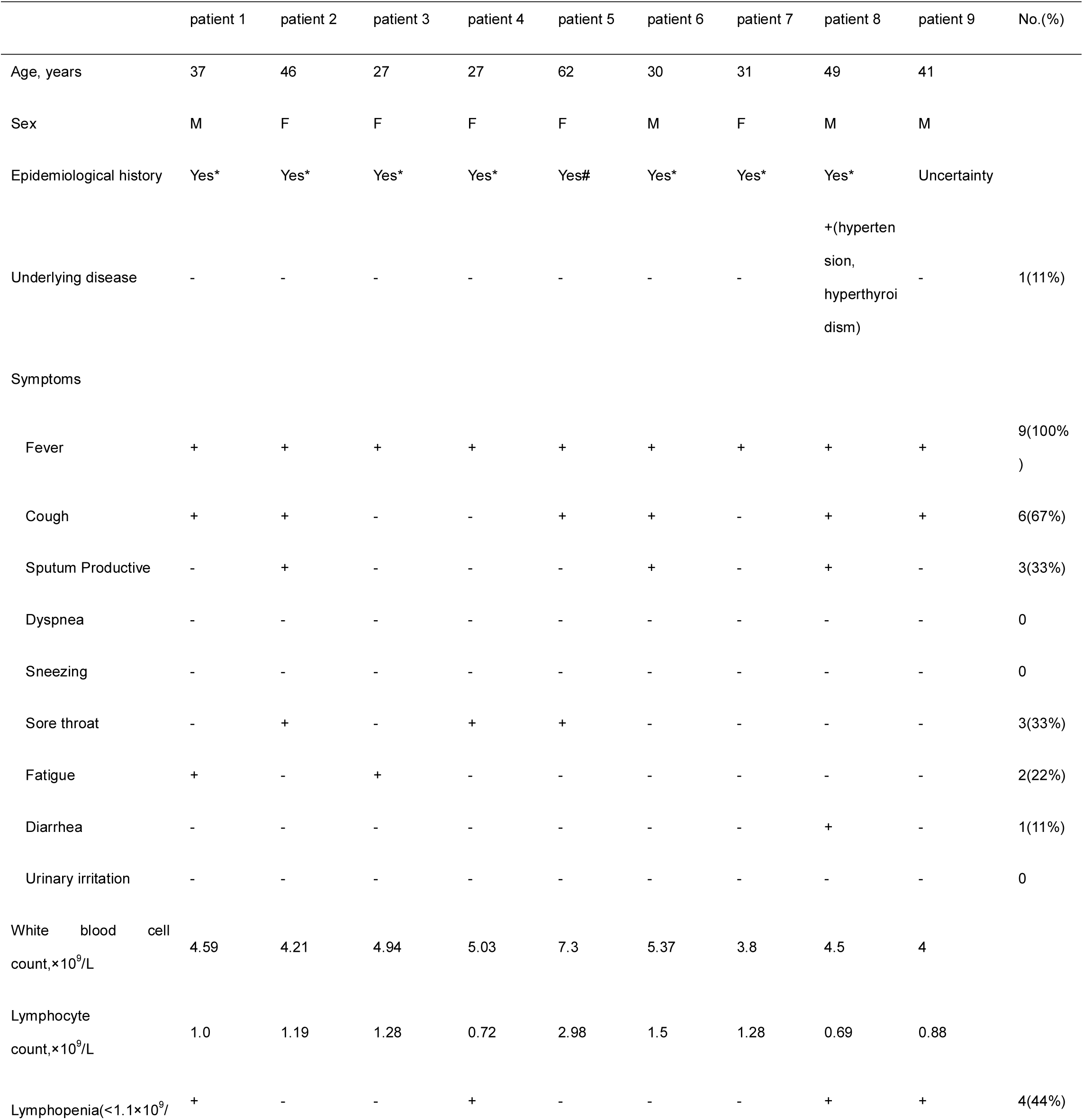
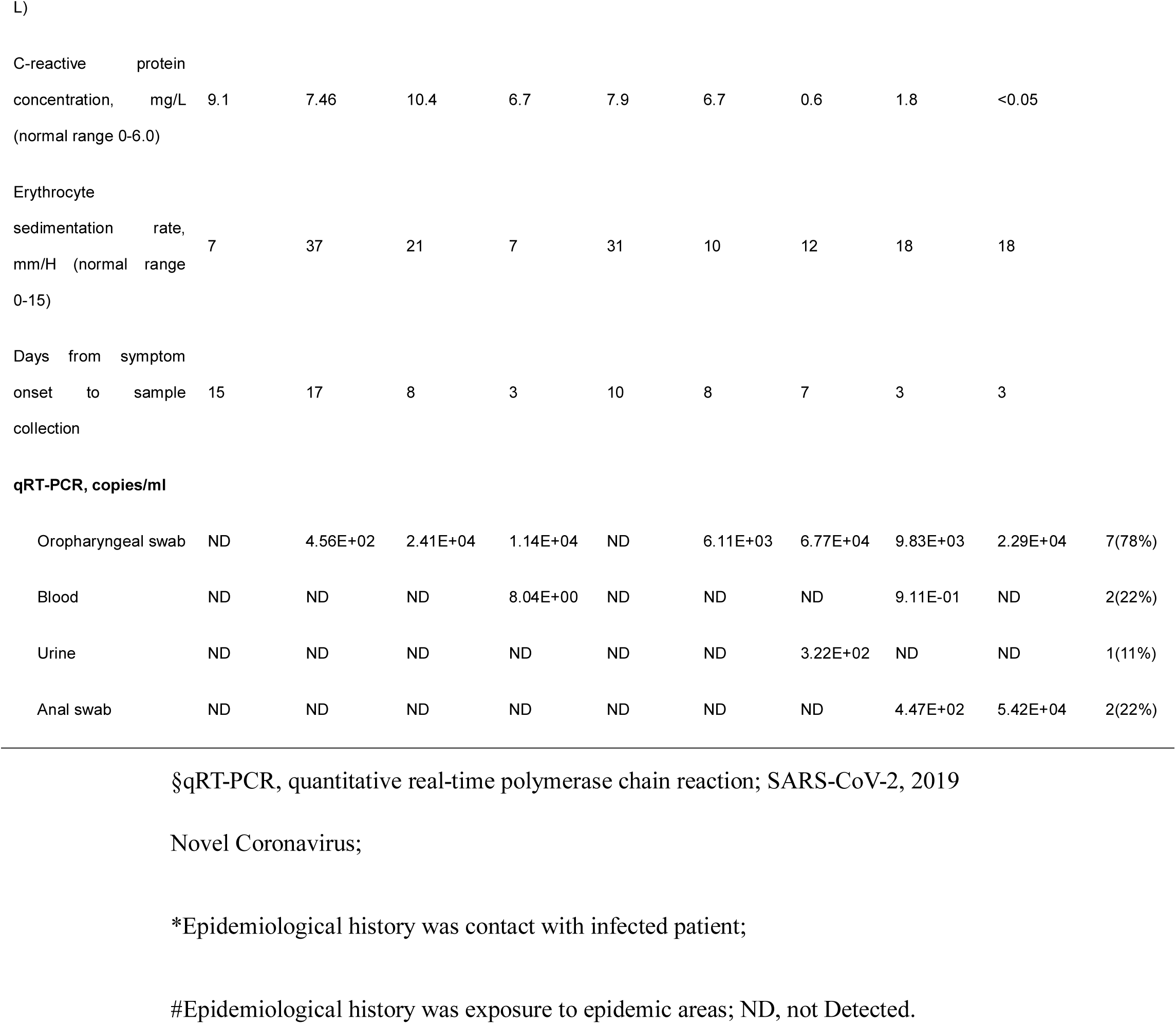
Clinical characteristics and qRT-PCR testing for the 4 kinds of specimens of patients infected with SARS-CoV-2§

|  | patient 1 | patient 2 | patient 3 | patient 4 | patient 5 | patient 6 | patient 7 | patient 8 | patient 9 | No.(%) |
| --- | --- | --- | --- | --- | --- | --- | --- | --- | --- | --- |
| Age, years | 37 | 46 | 27 | 27 | 62 | 30 | 31 | 49 | 41 |  |
| Sex | M | F | F | F | F | M | F | M | M |  |
| Epidemiological history | Yes* | Yes* | Yes* | Yes* | Yes# | Yes* | Yes* | Yes* | Uncertainty |  |
| Underlying disease | - | - | - | - | - | - | - | +(hyperten<br>sion,<br>hyperthyroi<br>dism) | - | 1(11%) |
| Symptoms |  |  |  |  |  |  |  |  |  |  |
| Fever | + | + | + | + | + | + | + | + | + | 9(100%) |
| Cough | + | + | - | - | + | + | - | + | + | 6(67%) |
| Sputum Productive | - | + | - | - | - | + | - | + | - | 3(33%) |
| Dyspnea | - | - | - | - | - | - | - | - | - | 0 |
| Sneezing | - | - | - | - | - | - | - | - | - | 0 |
| Sore throat | - | + | - | + | + | - | - | - | - | 3(33%) |
| Fatigue | + | - | + | - | - | - | - | - | - | 2(22%) |
| Diarrhea | - | - | - | - | - | - | - | + | - | 1(11%) |
| Urinary irritation | - | - | - | - | - | - | - | - | - | 0 |
| White blood cell<br>count,x10<sup>9</sup>/L | 4.59 | 4.21 | 4.94 | 5.03 | 7.3 | 5.37 | 3.8 | 4.5 | 4 |  |
| Lymphocyte<br>count,x10<sup>9</sup>/L | 1.0 | 1.19 | 1.28 | 0.72 | 2.98 | 1.5 | 1.28 | 0.69 | 0.88 |  |
| Lymphopenia(<1.1x10<sup>9</sup>/ | + | - | - | + | - | - | - | + | + | 4(44%) |

L)
|  |  |  |  |  |  |  |  |  |  |
| --- | --- | --- | --- | --- | --- | --- | --- | --- | --- |
| C-reactive protein concentration, mg/L | 9.1 | 7.46 | 10.4 | 6.7 | 7.9 | 6.7 | 0.6 | 1.8 | <0.05 |

|  |  |  |  |  |  |  |  |  |  |
| --- | --- | --- | --- | --- | --- | --- | --- | --- | --- |
| Erythrocyte sedimentation rate, mm/H (normal range 0-15) | 7 | 37 | 21 | 7 | 31 | 10 | 12 | 18 | 18 |
| Days from symptom onset to sample collection | 15 | 17 | 8 | 3 | 10 | 8 | 7 | 3 | 3 |
| **qRT-PCR, copies/ml** |  |  |  |  |  |  |  |  |  |  |
| Oropharyngeal swab | ND | 4.56E+02 | 2.41E+04 | 1.14E+04 | ND | 6.11E+03 | 6.77E+04 | 9.83E+03 | 2.29E+04 | 7(78%) |
| Blood | ND | ND | ND | 8.04E+00 | ND | ND | ND | 9.11E-01 | ND | 2(22%) |
| Urine | ND | ND | ND | ND | ND | ND | 3.22E+02 | ND | ND | 1(11%) |
| Anal swab | ND | ND | ND | ND | ND | ND | ND | 4.47E+02 | 5.42E+04 | 2(22%) |

§qRT-PCR, quantitative real-time polymerase chain reaction; SARS-CoV-2, 2019
Novel Coronavirus;
\*Epidemiological history was contact with infected patient;

Eight of nine patients had an obvious history of epidemiological exposure to SARS-CoV-2. All patients presented with a fever. Other symptoms were cough, sore throat, fatigue and diarrhea. Four patients had a lymphopenia. Elevated C-reactive protein and erythrocyte sedimentation rate were found in 6 and 5 cases respectively. All patients had normal serum levels of procalcitonin, alanine transaminase, creatinine, myocardial enzyme and arterial partial oxygen pressure.

## Discussion

The pathogenic mechanism of SARS-CoV-2 infection is still unclear. Current evidences indicate that it can invade multiple organ systems, including the respiratory system, digestive system and hematological system(*3-5*). But whether it can invade the urinary system has not been reported. In our study, urine, blood, anal swab and oropharyngeal swab from 9 patients were retested by qRT-PCR. It is the first time for the SARS-CoV-2 found in urine, though no urinary irritation was found. In addition, the quantity of the virus in the anal swab was closer to that of the oropharyngeal test, and those in the blood and urine were less than that of the oropharyngeal test, indicating that the “clearing effect” of the digestive tract on the virus is not obvious. Nevertheless, the relative symptoms, including diarrhea and urinary irritation did not happen to every patient with virus in anal swab and urine specimens. All samples were negative at the earliest time of the 10^th^ day after onset in that mild patient. Therefore, we believed that SARS-CoV-2 can invade the urinary system, hematological system and digestive system other than the respiratory system, not always with relative symptoms. Mild patients can be self-limiting. This will prompt clinicians to pay attention to the clinical manifestations of multiple systems, even if the corresponding clinical symptoms do not appear. In addition, it is necessary to carry out multiple examinations of various specimens to assess changes in disease and prognosis.

## Data Availability

I promise all data referred to in the manuscript are available.

